# Adherence to a Mediterranean diet and leisure-time physical activity are protective factors against the initiation of antidepressant, anxiolytic, antipsychotic, and antiseizure drug use in older adults: a cohort study

**DOI:** 10.1101/2024.10.04.24314883

**Authors:** Marta H. Hernandez, Eleonora Fornara, Camille Lassale, Olga Castañer, Ramón Estruch, Emilio Ros, Miguel Ángel Martínez-González, Dolores Corella, Nancy Babio, José Lapetra, Enrique Gómez-Gracia, Fernando Arós, Miquel Fiol, Lluis Serra-Majem, Antoni Riera-Mestre, Alfredo Gea, Carolina Ortega-Azorín, Andrés Díaz-López, Montserrat Fitó, Álvaro Hernáez

## Abstract

**Background:** We aimed to investigate the association between long-term adherence to MedDiet and LTPA levels and the initiation of antidepressant, anxiolytic, antipsychotic and antiseizure medication in older adults.

**Methods.:** We assessed the relationship between the cumulative average of MedDiet adherence and LTPA and the initiation of medications in the PREvención con DIeta MEDiterránea (PREDIMED) study. Sample sizes ranged from 5,940 for anxiolytics to 6,896 for antipsychotics. Associations between the cumulative average of MedDiet adherence (per each one-point increase in the adherence score) and LTPA (per each increase in 20 metabolic equivalents of task-minute/day [METs-min/day]) with drug initiation were studied by multivariable Cox regressions (covariates: age, sex, PREDIMED intervention group, recruitment site, education, smoking habit, BMI, alcohol, and energy intake; multiple comparisons: Bonferroni method). We explored non-linear exposure-outcome associations using smoothed cubic splines and the interaction among both exposures.

**Results:** One-point increases in MedDiet adherence score were associated with 23-28% less initiation of antidepressants (hazard ratio [HR] 0.72, 95% confidence interval [CI] 0.67-0.77), anxiolytics (HR 0.75, 95%CI 0.70-0.81), antipsychotics (HR 0.77, 95%CI 0.65-0.91), and antiseizure drugs (HR 0.77, 95%CI 0.69-0.85). Associations for anxiolytics and antiseizure drugs were particularly strong among participants with poor MedDiet adherence. Relationships between LTPA and initiation of antidepressant and anxiolytic drug use were non-linear. Among participants with the lowest LTPA values (0-150 METs-min/day), 20 METs-min/day increases were associated with 20% less risk of initiating antidepressant use (HR 0.80, 95%CI 0.75-0.86) and 15% less risk of initiating anxiolytic medication (HR 0.85, 95%CI 0.79-0.90). 20 METs-min/day increases were linearly associated with less initiation of antiseizure drugs (HR 0.96, 95%CI 0.94-0.99), and no clear associations were found for antipsychotic drugs. Individuals with high MedDiet adherence (≥10 adherence points) and high LTPA levels (≥150 METs-min/day) showed 42-59% less risk of initiating psychoactive drugs (antidepressants: HR 0.41, 95%CI 0.30-0.56; anxiolytics: HR 0.54, 95%CI 0.41-0.71; antipsychotics: HR 0.45, 95%CI 0.21-0.95; antiseizure drugs: HR 0.58, 95%CI 0.37-0.90). The combination was additive for antidepressants, antipsychotics and antiseizure drugs and synergistic for anxiolytics (*p*-interaction = 0.076).

**Conclusions:** Sustained adherence to MedDiet and LTPA were linked to lower initiation of psychoactive drugs in older adults.

## INTRODUCTION

Mental disorders represent a major burden for patients and society as they are leading causes of disability [1] and are associated with increased mortality rates [2,3]. This burden of morbidity and mortality is on the rise across all age, sex and socioeconomic levels [1,4]. The primary pharmacological treatments for these conditions typically involve antidepressants, anxiolytics, and, to a lesser extent, some antipsychotic and antiseizure medications. Older adults (≥65 years old) in European countries show a relevant prevalence of psychoactive drug use according to the latest National Health Surveys [5,6]. However, despite significant advancements, conventional treatments exhibit limited effectiveness and some adverse effects [7–9]. Older adults face greater risks when exposed to these drugs due to characteristics in this population such as changes in pharmacokinetics, polypharmacy, cognitive impairment, and frailty [10]. Consequently, patients and clinicians are increasingly interested in exploring complementary methods, such as adopting healthy dietary patterns or engaging in physical activity, to reduce the burden of mental disorders in the elderly population or, at least, delay and limit the initiation of psychoactive medication use [11,12].

Lifestyle psychiatry is a complex field due to the potential for reverse causation: an unhealthy lifestyle may increase the incidence of mental disorders and neurological conditions, which in turn are associated with a decline in dietary quality and reduced physical activity [13]. However, recent meta-analyses of observational data have provided evidence of the association between healthy diets and lower risk of depression [14] and of physical activity in warding off mental disorders [13]. Healthy dietary patterns such as the Mediterranean diet (MedDiet) and leisure-time physical activity (LTPA) have shown a protective potential on several risk factors for psychiatric and mental disorders such as oxidative stress, low-grade inflammation, gut microbiome composition, blood pressure, and metabolic indicators [15–17] [18–21].. Nevertheless, there is a scarcity of research investigating the association between MedDiet and LTPA and the incidence of clinical outcomes related to mental disorders such as the initiation of psychoactive medication use, using prospective study designs. Therefore, the main objective of this study is to investigate the association between long-term adherence to MedDiet and LTPA (individually and combined) and the initiation of psychoactive medication in older adults at high cardiovascular risk.

## MATERIALS AND METHODS

### Study population

This study is an observational, prospective analysis utilizing data from participants enrolled in the PREvención con DIeta MEDiterránea (PREDIMED) trial. The PREDIMED study was a multicenter, randomized controlled trial conducted in Spain from 2003 to 2010. Its principal objective was to assess the effects of a nutritional intervention based on a MedDiet supplemented with extra-virgin olive oil and mixed nuts, compared to a low-fat control diet, on the primary prevention of significant cardiovascular events in older adults at high cardiovascular risk. [22]. Eligible individuals were women (aged 60-80 years) and men (aged 55-80 years) without cardiovascular disease at enrolment but with type 2 diabetes or at least three of the following risk factors: hypertension (systolic blood pressure ≥140 mmHg, diastolic blood pressure ≥90 mmHg, use of antihypertensive drugs), high concentrations of low-density lipoprotein cholesterol (≥160 mg/dL), low levels of high-density lipoprotein cholesterol <50 mg/dL in women, (<40 mg/dL in men), body mass index >25 kg/m^2^, smoking, and family history of premature coronary disease (<55 years old in first degree male relatives, <65 years old in first degree female relatives).

For the present investigation, we used the PREDIMED data as an observational prospective cohort. We therefore adjusted all analyses for intervention groups. Of the 7,447 participants, we first excluded participants with no baseline data on MedDiet adherence (*n* = 22), no data from food frequency questionnaire (necessary for the estimated alcohol and energy intake, *n* = 39), outliers of the MedDiet adherence longitudinal average (*n* = 14), outliers of the LTPA longitudinal average (*n* = 39), and no medication use data (*n* = 328). Analyses of initiation of psychoactive drugs were performed in non-users at study baseline. Therefore, we also excluded users of each specific drug at baseline in the respective analyses. The study flow chart with the definitive sample sizes for all analyses is available in **Figure 1**. Our study is presented following the STrengthening the Reporting of Observational Studies in Epidemiology guidelines for reporting cohort studies.

**Figure 1.**
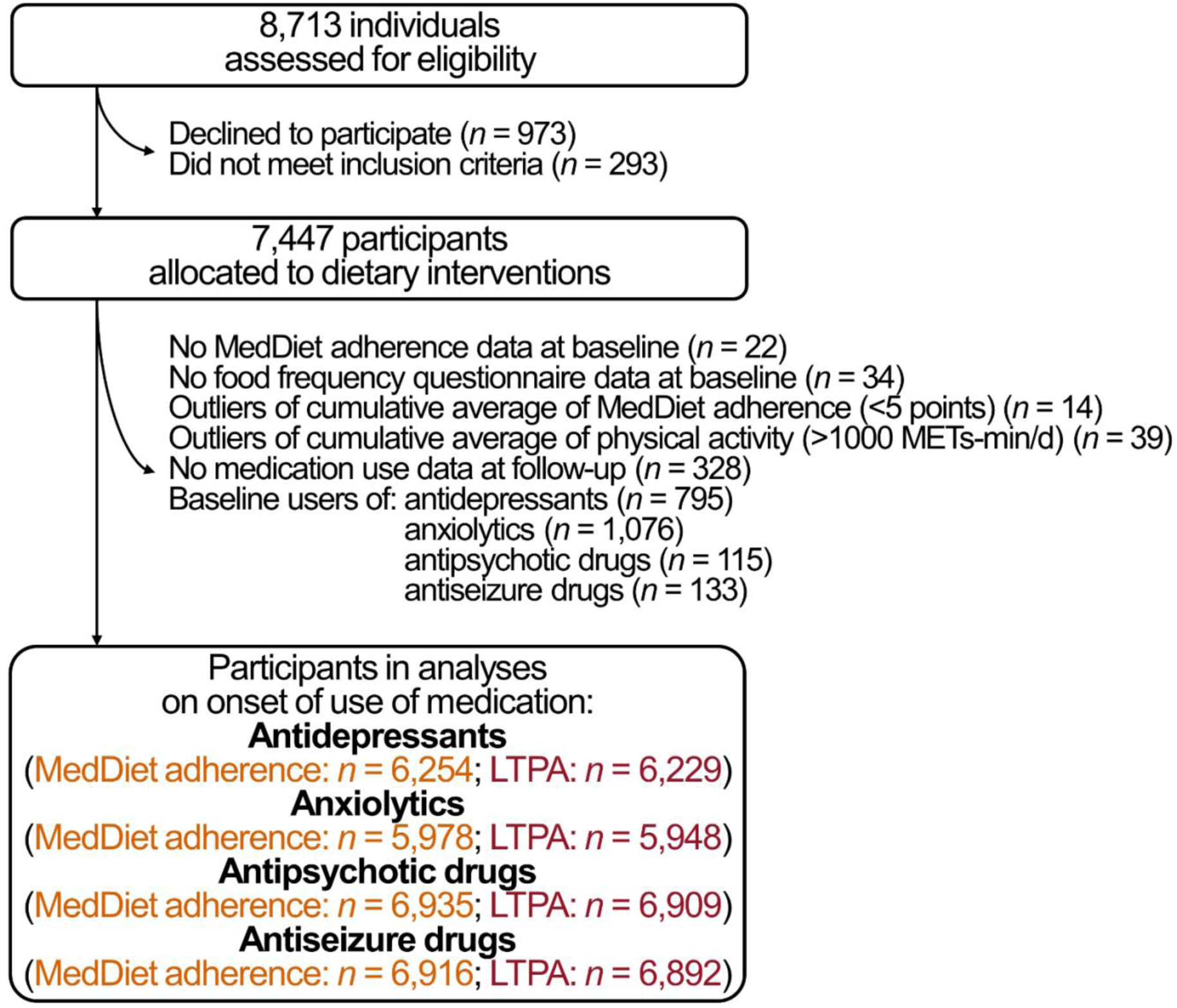
Flow chart.

### Exposures: adherence to a Mediterranean diet and leisure-time physical activity

At each visit, adherence to a MedDiet was assessed using the MedDiet adherence score. This score was based on a screener designed to evaluate the adherence to 14 dietary traits of the MedDiet, validated for Spanish adults [23]. The consumption of the following items contributed positively: (1) use of olive oil as main fat for cooking/seasoning; (2) ≥4 tablespoons/day of olive oil; (3) ≥3 servings/week of mixed nuts (30 g); (4) ≥2 servings/day of vegetables; (5) ≥3 servings/day of fruit; (6) ≥3 servings/week of legumes (150 g, boiled); (7) ≥3 servings/week of fish or seafood; (8) ≥2 servings/week of “sofrito”-based dishes (recipes with a base of stir-fried onion, garlic, tomato/pepper, and herbs cooked in olive oil); (9) wine in moderation (100 mL/day on average, within meals); (10) <1 serving/day of red and processed meat; (11) preference of poultry and rabbit over red and processed meat; (12) <1 serving/day of butter, margarine, or cream; (13) <1 carbonated or sugar-sweetened beverage/day; and (14) <2 servings/week of non-homemade pastries or sweets [23].

The estimation of LTPA was conducted using the Minnesota Leisure-Time Physical Activity Questionnaire, validated in both Spanish men and women [24,25]. This questionnaire captured information on the frequency and duration of sixty-seven activities performed by participants over the course of the previous year. LTPA was quantified in metabolic equivalents of task-minute per day (METs-min/d) by multiplying the metabolic equivalents of task associated with each activity by its average duration in minutes per day.

### Outcomes: initiation of use of psychoactive drugs

During the baseline and yearly follow-up examinations, information was gathered on medication use. Using the list of all the drugs that were considered in each family of psychoactive drugs available in Spain in 2003-2010 with their respecting Anatomical Therapeutic Chemical codes available in the **Supplemental Material**, we created variables of use (yes/no) of antidepressants, anxiolytics, antipsychotics, and antiseizure drugs. We defined the incidence of the initiation of use of any antidepressant, anxiolytic, antipsychotic, and antiseizure drug among those who were non-users at baseline. Initiation of use referred to the commencement of medication use that persisted for a minimum of three subsequent follow-up visits and drug use was unreported in only one visit between these three [26].

### Other variables

Trained professionals gathered initial data on various factors, including age (continuous), sex, educational attainment (primary/secondary/higher/unavailable), the prevalence of cardiovascular risk factors at baseline (diabetes, hypercholesterolemia, hypertriglyceridemia, and hypertension; yes/no), baseline body mass index (continuous), and baseline smoking status (never/former/current smoker) [22,27]. Using a validated 137-item semiquantitative food frequency questionnaire, we determined the alcohol consumption (in grams/day) and energy intake (in kilocalories/day) at baseline [27].

### Statistical analyses

The characteristics of the participants are described using means and standard deviations for normally distributed continuous variables, medians and interquartile ranges for non-normally distributed continuous variables, and proportions for categorical variables, overall and across groups defined by MedDiet adherence (low [<10 points] and high [≥10 points]) and by LTPA levels (an arbitrary threshold for low [<100 METs-min/day] and high levels [≥100 METs-min/day]).

We examined the relationships between the cumulative mean of MedDiet adherence or LTPA levels and the risk of initiating psychoactive medication use (as hazard ratios, HR) using Cox proportional hazards regression models. The cumulative mean of MedDiet adherence or LTPA was calculated as the average of all MedDiet adherence scores or LTPA values up until the occurrence of the outcome (incident cases) or the last available study visit (non-cases). Participants with outlier values in the cumulative mean of MedDiet adherence score (<5 points) or LTPA levels (>1000 METs-min/day) were excluded from the analysis. The reference cut-point was set at the minimum value for each exposure variable (5 points for MedDiet adherence score and 0 METs-min/day for LTPA). The follow-up time for each event was the time elapsed between the date of entry into the study and the date of the event or end of follow-up (December 1, 2010), whichever came first. As these are non-acute events, we assume that the occurrence of the event is dated at the midpoint between the last visit at which the volunteer was not treated with the medication and the first visit at which the treatment was registered[26,28,29]. The models were adjusted for sex, recruitment site, educational level (as strata variables), age, smoking habit, body mass index, alcohol consumption, energy intake, and intervention group. MedDiet adherence analyses were further adjusted for LTPA at baseline, while the LTPA analyses were further adjusted for MedDiet adherence score at baseline. To address intra-cluster correlations, robust variance estimators were used in all survival analyses considering as clusters the members of the same household [22]. Results were reported as HRs per each 1-point increase in MedDiet score and per each increase in 20 METs-min/day for LTPA (equivalent to increasing 40 minutes/week of brisk walking or Pilates, or 30 minutes/week of dancing or aerobic classes). We corrected our findings in linear analyses for multiple comparisons (two exposures × four outcomes = eight comparisons) by the Bonferroni method (*p*-value threshold < 0.00625).

We also examined the potential non-linear relationships between the cumulative mean of MedDiet adherence or LTPA levels and the risk of initiating psychoactive medication use. Initially, we explored whether a model using smoothed cubic splines (with *K*+4 degrees of freedom) to model the relationship between the exposure and the outcome. We used a likelihood ratio test to assess if this fitted the data better than a simple linear term. When the test was significant, we used smoothed cubic splines graphically present the associations [30]. We also reported the association between MedDiet adherence or LTPA and the risk of drug initiation at the lowest part of the curves (after graphic inspection, for participants with MedDiet adherence scores ranging from 5 to 8 points and LTPA levels ranging from 0 to 150 METs-min/day).

Additionally, we investigated whether the combination of high MedDiet adherence (≥10 points [31]) and high LTPA levels (≥150 MET-min/day, based on our findings, see below) was associated with a lower risk of drug use initiation, either additively or synergistically. Participants were classified into four groups: 1) low MedDiet adherence + low LTPA levels (reference group); 2) low MedDiet adherence + high LTPA levels; 3) high MedDiet adherence + low LTPA levels; and 4) high MedDiet adherence + high LTPA levels. We examined the difference in the risk of drug use initiation for groups 2, 3, and 4 compared to the reference group using Cox proportional hazards regression models. To assess potential synergy, we applied a likelihood ratio test comparing models with and without the interaction term for “MedDiet adherence × LTPA levels.” A significant interaction would suggest that the combination of the two exposures is associated with a stronger relationship than expected from their independent associations.

We performed the analyses using the “survival” package in R Software (version 4.3.1). Code for data management and analysis is available in https://github.com/alvarohernaez/MedDiet_LTPA_psychoactive_drugs.

## RESULTS

### Study population

The participants had an average age of 67 years, 58% of them were women, 14% we current smokers and –as per the selection criteria– they had a high prevalence of cardiovascular risk factors at baseline: 83% hypertension, 72% hypercholesterolemia, 49% diabetes, 47% obesity, 29% with hypertriglyceridemia (**Table 1**). Participants with a high adherence to a MedDiet and high LTPA levels were more likely to be men, to have higher education, lower prevalence of cardiovascular risk factors (such as diabetes, hypertriglyceridemia, hypertension, and obesity), and slightly higher intakes of energy and alcohol.

**Table 1.** Population description

|  | All participants | MedDiet adherence score<br>(cumulative average) |  |  | LTPA levels<br>(cumulative average) |  |  |
| --- | --- | --- | --- | --- | --- | --- | --- |
|  |  | Low | High | *p*-value | Low | High | *p*-value |
| Age, years<br>(mean ± SD) | 67.0<br>± 6.17 | 67.2<br>± 6.25 | 66.7<br>± 6.07) | 0.001 | 67.6<br>± 6.50) | 66.8<br>± 6.05) | <0.001 |
| Females<br>(*n*, %) | 4,080<br>(57.8%) | 2,277<br>(60.9%) | 1,803<br>(54.2%) | <0.001 | 1,277<br>(75.3%) | 2,803<br>(52.2%) | <0.001 |
| Education: |  |  |  | 0.001 |  |  | <0.001 |
| Primary<br>(*n*, %) | 5,367<br>(76.0%) | 2,895<br>(77.5%) | 2,472<br>(74.3%) |  | 1,344<br>(79.2%) | 4,023<br>(75.0%) |  |
| Secondary<br>(*n*, %) | 1,066<br>(15.1%) | 539<br>(14.4%) | 527<br>(15.8%) |  | 240<br>(14.1%) | 826<br>(15.4%) |  |
| Higher<br>(*n*, %) | 503<br>(7.12%) | 230<br>(6.16%) | 273<br>(8.21%) |  | 82<br>(4.83%) | 421<br>(7.85%) |  |
| Not available<br>(*n*, %) | 127<br>(1.80%) | 72<br>(1.93%) | 55<br>(1.65%) |  | 31<br>(1.83%) | 96<br>(1.79%) |  |
| Tobacco use: |  |  |  | 0.001 |  |  | <0.001 |
| Never smokers<br>(*n*, %) | 4,345<br>(61.5%) | 2,356<br>(63.1%) | 1,989<br>(59.8%) |  | 1,194<br>(70.4%) | 3,151<br>(58.7%) |  |
| Ex-smokers<br>(*n*, %) | 1,733<br>(24.5%) | 848<br>(22.7%) | 885<br>(26.6%) |  | 293<br>(17.3%) | 1,440<br>(26.8%) |  |
| Current smokers<br>(*n*, %) | 985<br>(13.9%) | 532<br>(14.2%) | 453<br>(13.6%) |  | 210<br>(12.4%) | 775<br>(14.4%) |  |
| Type-II diabetes<br>(*n*, %) | 3,442<br>(48.7%) | 1,925<br>(51.5%) | 1,517<br>(45.6%) | <0.001 | 862<br>(50.8%) | 2,580<br>(48.1%) | 0.055 |
| Hypercholesterolemia | 5,087 | 2,667 | 2,420 | 0.216 | 1,210 | 3,877 | 0.467 |
| (n, %) | (72.0%) | (71.4%) | (72.7%) |  | (71.3%) | (72.3%) |  |
| Hypertriglyceridemia | 2,045 | 1,181 | 864 |  | 556 | 1,489 |  |
| (n, %) | (29.0%) | (31.6%) | (26.0%) | <0.001 | (32.8%) | (27.7%) | <0.001 |
| Hypertension | 5,834 | 3,136 | 2,698 |  | 1,451 | 4,383 |  |
| (n, %) | (82.6%) | (83.9%) | (81.1%) | 0.002 | (85.5%) | (81.7%) | <0.001 |
| BMI categories: |  |  |  | <0.001 |  |  | <0.001 |
| <25.0 kg/m<sup>2</sup> | 523 | 237 | 286 |  | 90 | 433 |  |
| (n, %) | (7.40%) | (6.34%) | (8.60%) |  | (5.30%) | (8.07%) |  |
| 25.0-29.9 kg/m<sup>2</sup> | 3,207 | 1,564 | 1,643 |  | 583 | 2,624 |  |
| (n, %) | (45.4%) | (41.9%) | (49.4%) |  | (34.4%) | (48.9%) |  |
| ≥30.0 kg/m<sup>2</sup> | 3,333 | 1,935 | 1,398 |  | 1,024 | 2,309 |  |
| (n, %) | (47.2%) | (51.8%) | (42.0%) |  | (60.3%) | (43.0%) |  |
| Alcohol, g/d | 1.49 | 1.09 | 4.38 | <0.001 | 0.68 | 2.17 | <0.001 |
| (median, 1<sup>st</sup>-3<sup>rd</sup> quartile) | [0.00;10.4] | [0.00;8.84] | [0.00;12.4] |  | [0.00;5.92] | [0.00;11.7] |  |
| Energy, kcal/d | 2,274 | 2,201 | 2,356 | <0.001 | 2,239 | 2,285 | 0.009 |
| (mean ± SD) | ± 604 | ± 609 | ± 587 |  | ± 633 | ± 594 |  |

The analytical samples were as follows: for antidepressants, *n* = 6,215, with a median follow-up of 4.2 years and 7.9% became new users; for anxiolytics, *n* = 5,940, with a 4.2-year follow-up and 8.8% became new users; for antipsychotics, *n* = 6,896, with a 4.7-year follow-up and 1.1% became new users; for antiseizure medications, *n* = 6,878, with a 4.7-year follow-up and 2.8% became new users.

### MedDiet adherence and risk of psychoactive drug initiation

A one-point increase in cumulative MedDiet adherence was linearly associated with a 28% less risk of initiating antidepressant use (HR 0.72, 95% CI 0.67 to 0.78, **Figure 2A**), a 25% reduced risk of initiating anxiolytic medication (HR 0.75, 95% CI 0.70 to 0.81, **Figure 2B**), a 23% less risk of initiating antipsychotic use (HR 0.77, 95% 0.65 to 0.91, **Figure 2C**), and a 23% reduced risk of initiating antiseizure drugs (HR 0.77, 95% 0.69 to 0.85, **Figure 2D**). The risk of initiating anxiolytics, antipsychotics, and antiseizure drugs was better explained by a non-linear equation. There was a greater reduction in the risk of starting anxiolytics or antiseizure for each one-point MedDiet score increase in participants with low adherence scores (5-8 points) —a 55% lower risk for anxiolytics (**Figure 2B**) and a 43% lower risk for antiseizure drugs (**Figure 2D**). In the case of antipsychotics (**Figure 2C**), the association between greater MedDiet adherence and a lower risk of initiating such treatment was only observed among participants with greater adherence (8 points or more).

**Figure 2.**
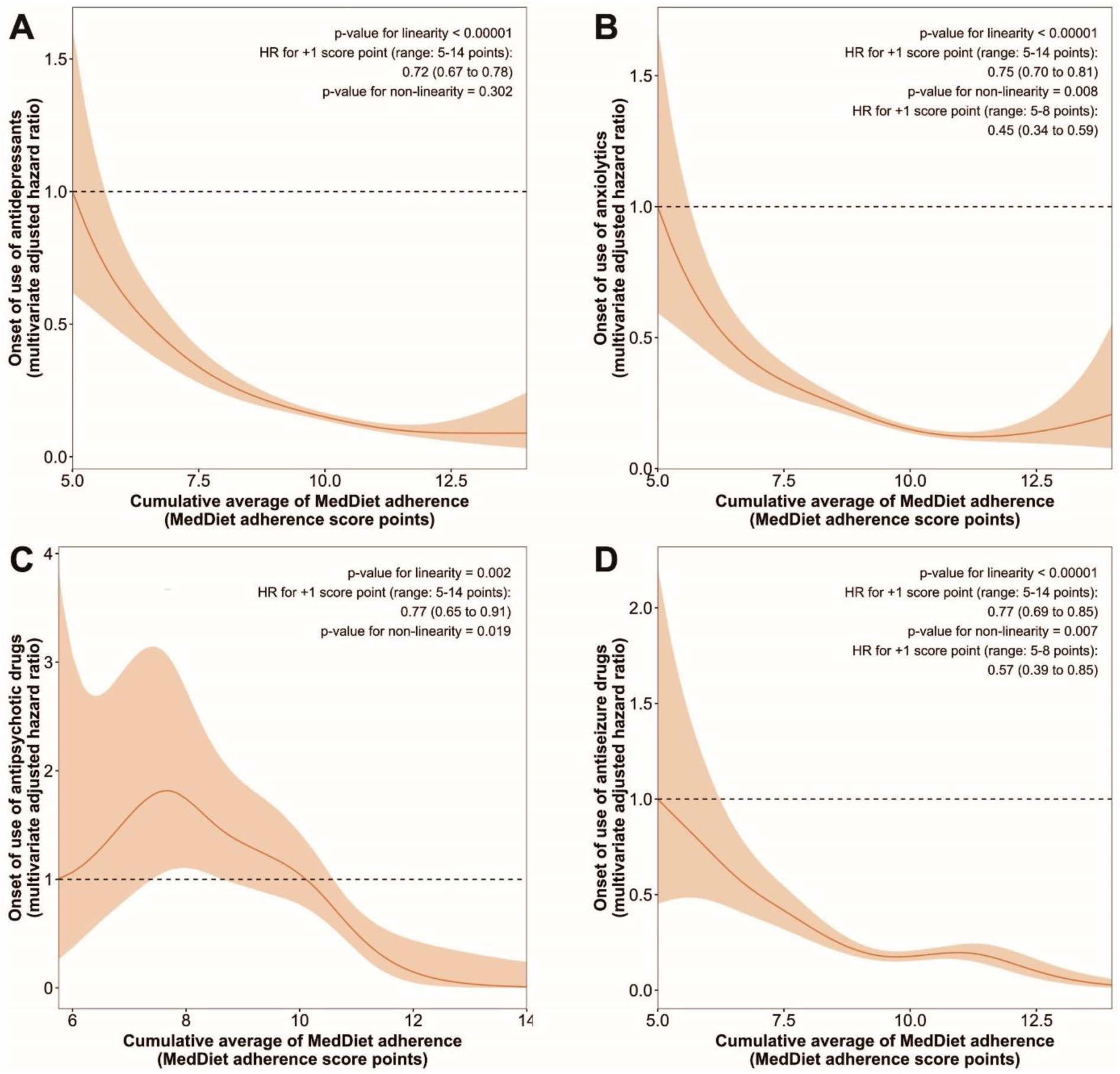
Hazards ratios and confidence intervals of the association between long-term adherence to a MedDiet and the risk of initiating the use of antidepressants (A), anxiolytics (B), antipsychotics (C) and antiseizure medication (D). Cox proportional hazards regression models with smoothed cubic splines were stratified by sex, recruitment site, and educational level, and adjusted for: age, smoking habit, body mass index, alcohol consumption, energy intake, leisure-time physical activity, and PREDIMED intervention group. Robust variance estimators were used to account for intra-cluster correlations.

### LTPA levels and risk of psychoactive drug initiation

A 20 METs-min/day increases in LTPA were linearly associated with a 3% reduced risk of initiating antidepressant use (HR 0.97, 95% CI 0.95 to 0.98, **Figure 3A**) and a 4% less risk of initiating antiseizure drugs (HR 0.96, 95% CI 0.94 to 0.99, **Figure 3D**), and nominally linked to a 2% reduced risk of initiating anxiolytic use (HR 0.98, 95% CI 0.97 to 1.00, **Figure 3B**). However, the risk of initiating antidepressant and anxiolytic drug use was better explained by non-linear equations. Among participants with the lowest LTPA values (0-150 METs-min/day), 20 METs-min/day increments in LTPA were associated with a 20% reduced risk of initiating antidepressant use (**Figure 3A**) and a 15% less risk of initiating anxiolytic medication (**Figure 3B**). No clear associations were found for antipsychotic drugs (**Figure 3C**).

**Figure 3.**
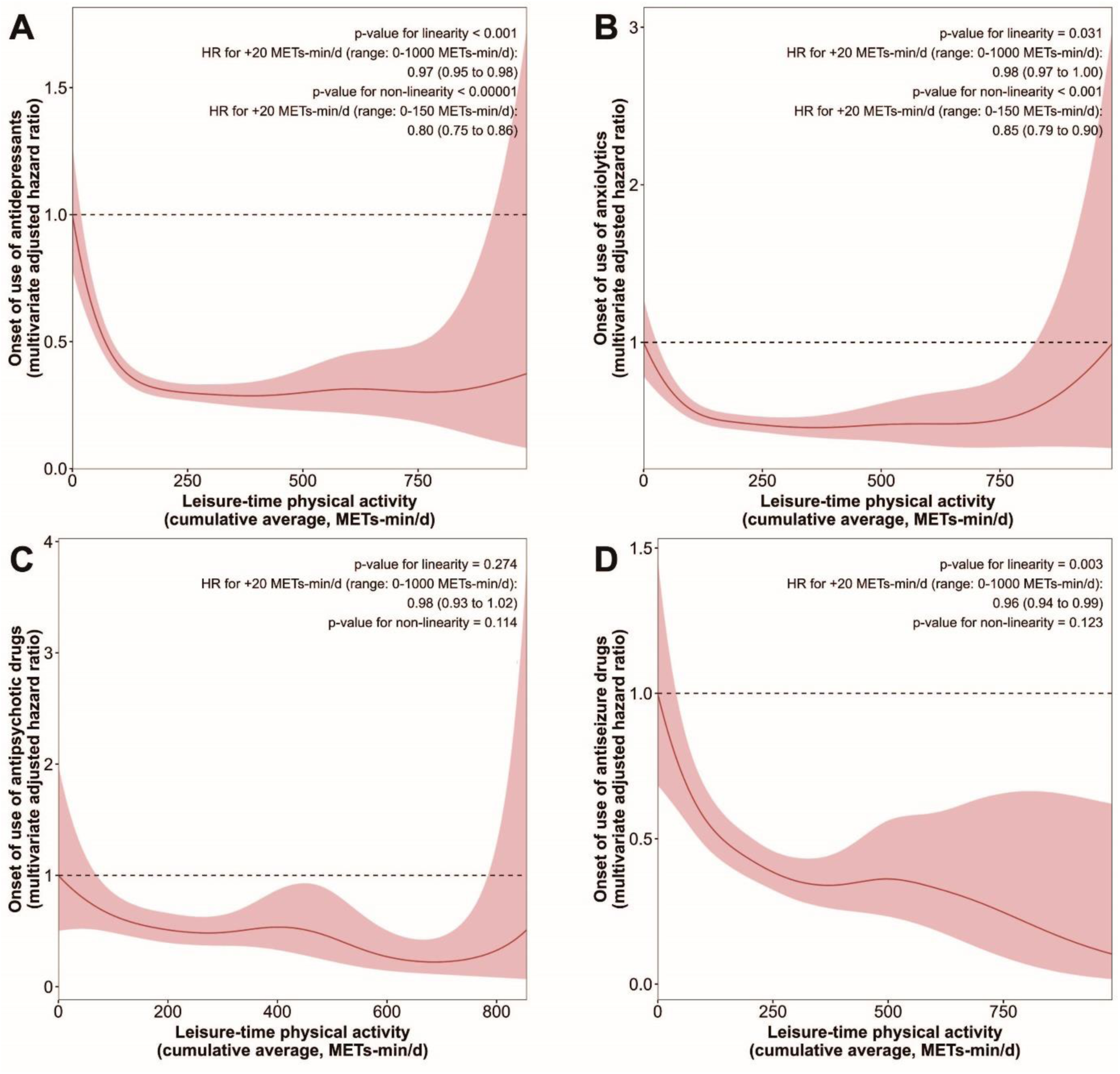
Association between long-term levels of leisure-time physical activity and the risk of initiating the use of antidepressants (A), anxiolytics (B), antipsychotics (C) and antiseizure drugs (D). Cox proportional hazards regression models with smoothed cubic splines were stratified by sex, recruitment site, and educational level, and adjusted for: age, smoking habit, body mass index, alcohol consumption, energy intake, adherence to a MedDiet, and PREDIMED intervention group. Robust variance estimators were used to account for intra-cluster correlations.

### Combination of MedDiet adherence and LTPA levels and risk of initiation of psychoactive drug use

High levels of MedDiet adherence (≥10 score points) and LTPA (≥150 METs-min/day) combined were associated with lower initiation of psychoactive therapy: 59% less risk of initiating antidepressant use (HR 0.41, 95% CI 0.30 to 0.56), 46% less risk of initiating anxiolytics (HR 0.54, 95% CI 0.41 to 0.71), 55% less risk of initiating antipsychotics (HR 0.45, 95% CI 0.21 to 0.95) and 41% less risk of initiating antiseizure therapy (HR 0.58, 95% CI 0.37 to 0.90) (**Figure 4**). The magnitude of the association of high MedDiet and high LTPA combined was synergistic for anxiolytics (*p*-value for interaction = 0.076) and additive for antidepressants, antipsychotics, and antiseizure medications.

**Figure 4.**
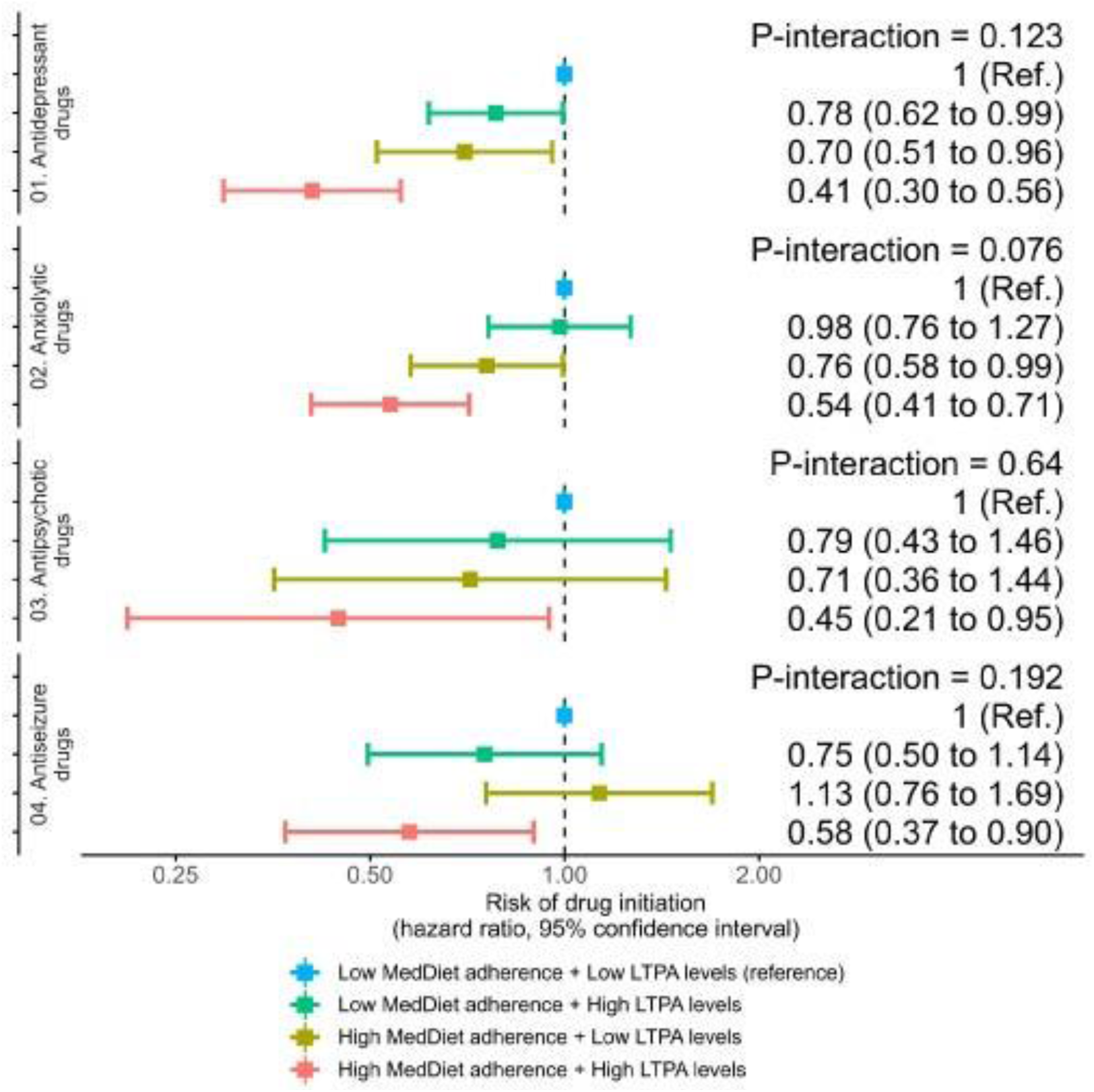
Levels of MedDiet adherence and leisure-time physical activity and risk of initiating psychoactive drugs.

## DISCUSION

Among older adults at high cardiovascular risk, we found that adherence to a MedDiet was associated with a lower risk of initiating the use of antidepressants, anxiolytics, antipsychotics, and antiseizure drugs. In the case of anxiolytics and antiseizure drugs, the association was particularly strong among participants with poor sustained adherence to a MedDiet. Our findings also indicate a strong association between slight increases in LTPA (equivalent to 40 minutes/week of brisk walking or Pilates, or 30 minutes/week of dancing or aerobic classes) and lower risk of initiating the use of antidepressants, anxiolytics and antiseizure medication. The magnitudes of the associations were stronger at lower levels of LTPA. Furthermore, individuals with high levels of both exposures show a particularly strong association with lower risk of initiation of psychoactive drugs, which was synergistic for anxiolytic medication.

Previous studies have highlighted the neuroprotective properties of a traditional MedDiet on mental health. These benefits could be due to its capacity to reduce oxidative stress and low-grade inflammation thanks to its array of bioactive compounds, including antioxidants, polyunsaturated and monounsaturated fatty acids, and prebiotic fiber [18,32,33]. Our results show an association between higher adherence to a MedDiet and a lower incidence of initiating antidepressant, anxiolytic, antipsychotic, and antiseizure medications in older adults, with a particularly pronounced association for anxiolytics and antiseizure drugs among those with very low MedDiet adherence sustained in time. High adherence to a MedDiet has already been linked to lower incident depression [14]. However, to the best of our knowledge, our study is the first to reveal that the MedDiet is linked to a reduced incidence of clinical outcomes related to anxiety, psychosis, and epilepsy. Prior research in this area includes only two cross-sectional studies that inversely correlated the MedDiet with anxiety symptoms in elderly individuals [34,35] and one cross-sectional study that examined the relationship between diet quality and schizophrenia [36], and no studies that have investigated the relationship of MedDiet with any clinical manifestation of epilepsy in adults.

Our results have also shown a link between increases in LTPA and a reduced incidence of initiating antidepressant and anxiolytic medications among participants with very low LTPA levels, as well as a relationship with lower initiation of antiseizure medication. Our findings are consistent with previous evidence, as exercise interventions have been shown to effectively treat symptoms of depression [37], anxiety [38], and epilepsy [39]. Our results also suggested a trend towards a lower incidence of initiating antipsychotic medication. Although this trend was not entirely significant, likely due to the low incidence rate in our population (1.1%), it aligns with previous evidence indicating that physical activity is beneficial for improving symptoms of psychotic disorders [40]. However, it should be noted that our results extend beyond previous evidence by showing that modest increases in physical activity—equivalent to a weekly session of 30-50 minutes of mild to moderate intensity exercise—are linked to a lower risk of starting antidepressant or anxiolytic medication, especially in those significantly below the recommended LTPA levels.

Our findings also suggest a synergistic association between a high adherence to MedDiet diet combined with high levels of LTPA and the initiation of anxiolytic drugs, as well as an additive relationship with the initiation of antidepressants, antipsychotics, and antiepileptic drugs. A healthy diet combined with physical activity has been traditionally linked to decreased mortality and lower incidence of chronic cardiometabolic disease [41,42], and their combination has been associated with reduced initiation of use of cardiovascular drugs in older adults [26]. Additionally, this combination has been linked to enhanced mental wellbeing [43], particularly in terms of depression [44] and improved symptoms of psychosis [45]. However, to the best of our knowledge, this is the first study to report similar findings in relation to mental health medications and a potentially synergistic benefit regarding anxiety.

We acknowledge some limitations in our study. Firstly, our analyses were based on data from the PREDIMED trial, which primarily focused on dietary intervention and did not include specific advice on LTPA. As a result, we treated the data as a cohort study, adjusting all our analyses for the intervention group. Nevertheless, the use of cumulative averages of MedDiet adherence and LTPA levels as exposure variables, instead of a single baseline measurement, is a strength of the study. Secondly, due to the available data, we could only collect categorical information regarding drug use, preventing us from studying dose changes or other aspects of medication use. Thirdly, the initiation of psychoactive-related therapy was not a predetermined endpoint in the PREDIMED study. Therefore, there was no standardization of treatment and patients were managed according to the clinical practice of the attending physicians and participating hospitals. Fourthly, main exposures and certain covariates, such as alcohol and energy intake, relied on self-reported data, which introduces the possibility of misclassification and potential bias. Lastly, our conclusions are applicable specifically to older adults at high cardiovascular risk and may not be generalizable to other populations.

## CONCLUSIONS

In a cohort of older adults at high cardiovascular risk, our results showed that both higher adherence to a MedDiet and LTPA levels were associated with a lower incidence of initiating common psychoactive medication, individually and combined. These findings underscore the potential of a healthy diet and regular physical activity to improve mental health and neurological outcomes among older individuals. Our work sets the stage for future randomized controlled trials to assess the effectiveness of MedDiet and physical activity interventions in preventing the onset of depression, anxiety, psychotic disorders, and seizures, particularly given the prevalence and disabling nature of these conditions in older adults, along with their associated economic burden on health systems.

## ETHICS APPROVAL AND CONSENT TO PARTICIPATE

The study protocol of the PREDIMED study complied with the Declaration of Helsinki for Medical Research involving Human Subjects. It received approval from institutional ethic committees at all recruitment sites. The protocol of this sub-study received approval from the Hospital del Mar Ethics Committee (Barcelona, Spain). The PREDIMED study protocol was registered with the International Standard Randomized Controlled Trial Number ISRCTN35739639 (http://www.isrctn.com/ISRCTN35739639) and is available in the PREDIMED study website (http://www.predimed.es). All participants provided written informed consent before joining the study.

## CONSENT FOR PUBLICATION

Not applicable.

## AVAILABILITY OF DATA AND MATERIALS

The dataset analyzed during the current study cannot be made publicly available due to national data regulations and ethical considerations, including the absence of explicit written consent from study participants to make their deidentified data available upon study completion. However, data described in the manuscript will be provided to bona fide investigators for collaboration upon request and approval. Requests can be made by sending a letter to the PREDIMED Steering Committee.

## COMPETING INTERESTS

R.E. reports being a board member of the Research Foundation on Wine and Nutrition, the Beer and Health Foundation, and the European Foundation for Alcohol Research; personal fees from KAO Corporation; lecture fees from Instituto Cervantes, Fundación Dieta Mediterranea, Cerveceros de España, Lilly Laboratories, AstraZeneca, and Sanofi; and grants from Novartis, Amgen, Bicentury, and Grand Fountaine. E.R. reports personal fees, grants, and nonfinancial support from the California Walnut Commission and Alexion; and nonfinancial support from the International Nut Council. J.S.-S. reports being a board member and personal fees from Instituto Danone Spain; being a board member and grants from the International Nut and Dried Fruit Foundation. The rest of the authors have nothing to disclose.

## FUNDING

This work was supported by the Blanquerna School of Health Sciences (University Ramon Llull, Barcelona, Spain) and the Instituto de Salud Carlos III [grant number CD17/00122]. The funders had no role in the study design; the collection, analysis, and interpretation of data; in the writing of the report; or in the decision to submit the article for publication. The views and opinions expressed in this paper are those of the authors only and do not necessarily reflect those of the funders.

## AUTHORS’ CONTRIBUTIONS

M.H.H. was responsible for formal analyses, data interpretation, visualization, and drafting the article. E.F. contributed to data interpretation and revised the article critically. C.L. contributed to data interpretation and revised the article critically. O.C. contributed to data interpretation and revised the article critically. R.E. contributed to funding and resource acquisition, investigation, project administration, data interpretation, and revised the article critically. E.R. contributed to funding and resource acquisition, investigation, project administration, data interpretation, and revised the article critically. M.Á.M.-G. contributed to funding and resource acquisition, investigation, project administration, data interpretation, and revised the article critically. D.C. contributed to funding and resource acquisition, investigation, project administration, data interpretation, and revised the article critically. N.B. contributed to data interpretation and revised the article critically. J.L. contributed to data interpretation and revised the article critically. E.G.-G. contributed to data interpretation and revised the article critically.

F.A. contributed to data interpretation and revised the article critically. M.Fiol contributed to data interpretation and revised the article critically. L.S.-M. contributed to data interpretation and revised the article critically. A.R.M contributed to data interpretation and revised the article critically. A.G. contributed to data interpretation and revised the article critically. C.O.-A. contributed to data interpretation and revised the article critically. A.D.-L. contributed to data interpretation and revised the article critically. M.Fitó contributed to funding and resource acquisition, investigation, project administration, data interpretation, and revised the article critically. A.H. coordinated the project, conceived and designed the study, contributed to funding acquisition, was responsible for data curation, contributed to data interpretation, and critically revised the article. M.H.H. and A.H. are the guarantors of this study, accept full responsibility for the work and the conduct of the study, have access to the data, and controlled the decision to publish. All authors read and approved the final manuscript.

## ACKNOWLEDGEMENTS

We are extremely grateful to the PREDIMED Study participants for their collaboration. A full list of the names of all the PREDIMED Study collaborators is available in the **Appendix**. CIBER de Fisiopatología de la Obesidad y Nutrición (CIBEROBN), CIBER de Epidemiología y Salud Pública (CIBERESP) and CIBER de Enfermedades Cardiovasculares (CIBERCV) are initiatives of Instituto de Salud Carlos III (Madrid, Spain), and are financed by the European Regional Development Fund.

## ABBREVIATIONS

HR: hazard ratio
LTPA: leisure-time physical activity
MedDiet: Mediterranean Diet
METs-min/day: metabolic equivalents of task per minute per day
PREDIMED: PREvención con DIeta MEDiterránea

## SUPPLEMENTAL MATERIALS

### Detailed list of antidepressants, anxiolytics, antipsychotics, and antiseizure drugs assessed in the study

Antidepressants: agomelatine (Anatomical Therapeutic Chemical code: N06AX22), amitriptyline (N06AA09), bupropion (N06AX12), citalopram (N06AB04), clomipramine (N06AA04), desipramine (N06AA01), desvenlafaxine (N06AX23), doxepin (N06AA12), dosulepin (N06AA16), duloxetine (N06AX21), escitalopram (N06AB10), fluoxetine (N06AB03), imipramin (N06AA02), maprotiline (N06AA21), mianserin (N06AX03), mirtazapine (N06AX11), moclobemide (N06AG02), nortriptyline (N06AA10), oxitriptan (N06AX01), paroxetine (N06AB05), reboxetine (N06AX18), sertraline (N06AB06), trazodone (N06AX05), and venlafaxine (N06AX16).

Anxiolytics: alprazolam (N05BA12), bentazepam (N05BA24), bromazepam (N05BA08), brotizolam (N05CD09), buspirone (N05BE01), clobazam (N05BA09), clonazepam (N03AE01), clordiazepoxide (N05BA02), clorazepate (N05BA05), diazepam (N05BA01), flunitrazepam (N05CD03), flurazepam (N05CD01), halazepam (N05BA13), ketazolam (N05BA10), lorazepam (N05BA06), lormetazepam (N05CD06), loprazolam (N05CD11), medazepam (N05BA03), midazolam (N05CD08), pinazepam (N05BA14), tepazepam (N05BA01), triazolam (N05CD05), zaleplon (N05CF03), zolpidem (N05CF02), and zopiclone (N05CF01).

Antipsychotics: amisulpride (N05AL05), chlorpromazine (N05AA01), clotiapine (N05AH06), flupentixol (N05AF01), fluphenazine (N05AB02), haloperidol (N05AD01), levomepromazine (N05AA02), olanzapine (N05AH03), perphenazine (N05AB03), pimozide (N05AG02), quetiapine (N05AH04), risperidone (N05AX08), sulpiride (N05AL01), thioridazine (N05AC02), tiapride (N05AL03), and ziprasidone (N05AE04).

Antiseizure drugs: carbamazepine (N03AF01), clonazepam (N03AE01), gabapentin (N02BF01), lamotrigine (N03AX09), levetiracetam (N03AX14), oxcarbazepine (N03AF02), phenobarbital (N03AA02), phenytoin (N03AB02), pregabalin (N02BF02), primidone (N03AA03), tiagabine (N03AG06), topiramate (N03AX11), valproic acid (N03AG01), valpromide (N03AG02), and zonisamide (N03AX15).

## APPENDIX

### Full list of PREDIMED study collaborators

Hospital Clinic, Institut d’Investigacions Biomèdiques August Pi i Sunyer, Barcelona, Spain: R. Estruch, M. Serra, A. Pérez-Heras, C. Viñas, R. Casas, L. de Santamaría, S. Romero, E. Sacanella, G. Chiva, P. Valderas, S. Arranz, J.M. Baena, M. García, M. Oller, J. Amat, I. Duaso, Y. García, C. Iglesias, C. Simón, L. Quinzavos, L. Parra, M. Liroz, J. Benavent, J. Clos, I. Pla, M. Amorós, M.T. Bonet, M.T. Martin, M.S. Sánchez, J. Altirriba, E. Manzano, A. Altés, M. Cofán, C. Valls-Pedret, A. Sala-Vila, M. Doménech, R. Gilabert, and N. Bargalló. University of Navarra, Primary Care Centres, Pamplona, Spain: M.Á. Martínez-González, A. Sánchez-Tainta, B. Sanjulián, E. Toledo, M. Bes-Rastrollo, A. Martí, C. Razquin, P. Buil-Cosiales, M. Serrano-Martínez, J. Díez-Espino, A. García-Arellano, I. Zazpe, F.J. Basterra-Gortari, E.H. Martínez-Lapiscina, A. Gea, M. Garcia-Lopez, J.M. Nuñez-Córdoba, N. Ortuño, N. Berrade, V. Extremera-Urabayen, C. Arroyo-Azpa, L García-Pérez, J. Villanueva-Tellería, F. Cortés-Ugalde, T. Sagredo-Arce, Mª D. García de la Noceda-Montoy, Mª D. Vigata-López, Mª T. Arceiz-Campo, A. Urtasun-Samper, Mª V. Gueto-Rubio, and B. Churio-Beraza. University of Valencia, Valencia, Spain; Universitat Jaume I, Castellon, Spain; and Conselleria de Sanitat, Generalitat Valenciana: D. Corella, P. Carrasco, C. Ortega-Azorín, E.M. Asensio, R. Osma, R. Barragán, F. Francés, M. Guillén, J.I. González, C. Sáiz, O. Portolés, F.J. Giménez, O. Coltell (U. Jaume I), R. Fernández-Carrión, I. González-Monje, L. Quiles, V. Pascual, C. Riera, M.A. Pages, D. Godoy, A. Carratalá-Calvo, S. Sánchez-Navarro, and C. Valero-Barceló.

University Rovira i Virgili, Reus, Spain: J. Salas-Salvadó, M. Bulló, R. González, C. Molina, F. Márquez, N. Babio, M. Sorlí, J. García-Roselló, F. Martin, R. Tort, A. Isach, B. Costa, J.J. Cabré, J. Fernández-Ballart, N. Ibarrola, C. Alegret, P. Martínez, S. Millán, J.L. Piñol, J. Basora, and J.M. Hernández.

Institut Hospital del Mar d’Investigacions Mèdiques, Barcelona, Spain: M. Fitó, M.I. Covas, O. Castañer, S. Tello, J. Vila, H. Schröder, R. De la Torre, D. Muñoz-Aguayo, N. Molina, E. Maestre, A. Rovira, R. Elosua, and M. Farré.

University Hospital of Alava, Vitoria, Spain: F. Arós, I. Salaverría, T. del Hierro, J. Algorta, S. Francisco, A. Alonso-Gómez, J. San-Vicente, E. Sanz, I. Felipe, A. Alonso-Gómez, and A. Loma-Osorio.

University of Málaga, Málaga, Spain: E. Gómez-Gracia, R. Benítez-Pont, M. Bianchi-Alba, J. Fernández-Crehuet Navajas, J. Wärnberg, R. Gómez-Huelgas, J. Martínez-González, V. Velasco-García, J. de Diego-Salas, A. Baca-Osorio, J. Gil-Zarzosa, J.J. Sánchez-Luque, and E. Vargas-López.

Institute of Health Sciences, University of Balearic Islands, and Hospital Son Espases, Palma de Mallorca, Spain: M. Fiol, M. García-Valdueza, M. Moñino, A. Proenza, R. Prieto, G. Frontera, M. Ginard, F. Fiol, A. Jover, and J. García.

Department of Family Medicine, Primary Care Division of Sevilla, Sevilla, Spain: J. Lapetra, M. Leal, E. Martínez, J.M. Santos, M. Ortega-Calvo, P. Román, F.J. García, P. Iglesias, Y. Corchado, E. Mayoral, and C. Lama.

University of Las Palmas de Gran Canaria, Las Palmas, Spain: L. Serra-Majem, J. Álvarez-Pérez, E. Díez-Benítez, I. Bautista-Castaño, I. Maldonado-Díaz, A. Sánchez-Villegas, F. Sarmiendo-de la Fe, C. Simón-García, I. Falcón-Sanabria, B. Macías-Gutiérrez, and A.J. Santana-Santana.

Hospital Universitario de Bellvitge, Hospitalet de Llobregat, Barcelona, Spain: X. Pintó, E. de la Cruz, A. Galera, Y. Soler, F. Trias, I. Sarasa, E. Padres, R. Figueras, X. Solanich, R. Pujol and E. Corbella.

Clinical End Point Committee: F. Arós (chair), M. Aldamiz, A. Alonso-Gómez, J. Berjón, L. Forga, J. Gállego, M. A. García-Layana, A. Larrauri, J. Portu, J. Timiraus, and M. Serrano-Martínez.

## Notes

### Competing Interest Statement

The authors have declared no competing interest.

## REFERENCES

1. GBD 2019 Mental Disorders Collaborators. Global, regional, and national burden of 12 mental disorders in 204 countries and territories, 1990-2019: a systematic analysis for the Global Burden of Disease Study 2019. Lancet Psychiatry. 2022;9:137–50.

2. Rehm J, Shield KD. Global Burden of Disease and the Impact of Mental and Addictive Disorders. Curr Psychiatry Rep. 2019;21:10.

3. Charlson FJ, Baxter AJ, Dua T, Degenhardt L, Whiteford HA, Vos T. Excess mortality from mental, neurological and substance use disorders in the Global Burden of Disease Study 2010. Epidemiol Psychiatr Sci. 2015;24:121–40.

4. Patel V, Saxena S, Lund C, Thornicroft G, Baingana F, Bolton P, et al. The Lancet Commission on global mental health and sustainable development. The Lancet. 2018;392:1553–98.

5. Maestre-Miquel C, López-de-Andrés A, Ji Z, de Miguel-Diez J, Brocate A, Sanz-Rojo S, et al. Gender Differences in the Prevalence of Mental Health, Psychological Distress and Psychotropic Medication Consumption in Spain: A Nationwide Population-Based Study. Int J Environ Res Public Health. 2021;18:6350.

6. Carrasco-Garrido P, López de Andrés A, Hernández Barrera V, Jiménez-Trujillo I, Jiménez-García R. National trends (2003-2009) and factors related to psychotropic medication use in community-dwelling elderly population. Int Psychogeriatr. 2013;25:328–38.

7. Van Zoonen K, Buntrock C, Ebert DD, Smit F, Reynolds CF, Beekman AT, et al. Preventing the onset of major depressive disorder: A meta-analytic review of psychological interventions. Int J Epidemiol. 2014;43:318–29.

8. Carbon M, Correll CU. Clinical predictors of therapeutic response to antipsychotics in schizophrenia. Dialogues Clin Neurosci. 2014;16:505–24.

9. Yu Z, Jiang H, Shao L, Zhou Y, Shi H, Ruan B. Use of antipsychotics and risk of myocardial infarction: a systematic review and meta-analysis. Br J Clin Pharmacol. 2016;82:624–32.

10. Loggia G, Attoh-Mensah E, Pothier K, Morello R, Lescure P, Bocca M-L, et al. Psychotropic Polypharmacy in Adults 55 Years or Older: A Risk for Impaired Global Cognition, Executive Function, and Mobility. Front Pharmacol. 2020;10:1659.

11. Bastos AA, Nogueira LR, Neto JV, Fisberg RM, Yannakoulia M, Ribeiro SML. Association between the adherence to the Mediterranean dietary pattern and common mental disorders among community-dwelling elders: 2015 Health Survey of São Paulo, SP, Brazil. J Affect Disord. 2020;265:389–94.

12. Mortazavi SS, Mohammad K, Ardebili HE, Beni RD, Mahmoodi M, Keshteli AH. Mental disorder prevention and physical activity in Iranian elderly. Int J Prev Med. 2012;3:S64–72.

13. Firth J, Solmi M, Wootton RE, Vancampfort D, Schuch FB, Hoare E, et al. A meta-review of “lifestyle psychiatry”: the role of exercise, smoking, diet and sleep in the prevention and treatment of mental disorders. World Psychiatry Off J World Psychiatr Assoc WPA. 2020;19:360–80.

14. Lassale C, Batty GD, Baghdadli A, Jacka F, Sánchez-Villegas A, Kivimäki M, et al. Healthy dietary indices and risk of depressive outcomes: a systematic review and meta-analysis of observational studies. Mol Psychiatry. 2019;24:965–86.

15. Dinu M, Pagliai G, Casini A, Sofi F. Mediterranean diet and multiple health outcomes: an umbrella review of meta-analyses of observational studies and randomised trials. Eur J Clin Nutr. 2018;72:30–43.

16. Pattyn N, Cornelissen VA, Eshghi SRT, Vanhees L. The Effect of Exercise on the Cardiovascular Risk Factors Constituting the Metabolic Syndrome: A Meta-Analysis of Controlled Trials. Sports Med. 2013;43:121–33.

17. Clark JS, Simpson BS, Murphy KJ. The role of a Mediterranean diet and physical activity in decreasing age-related inflammation through modulation of the gut microbiota composition. Br J Nutr. 2022;128:1299–314.

18. Marx W, Moseley G, Berk M, Jacka F. Nutritional psychiatry: the present state of the evidence. Proc Nutr Soc. 2017;76:427–36.

19. Bauer ME, Teixeira AL. Inflammation in psychiatric disorders: what comes first? Ann N Y Acad Sci. 2019;1437:57–67.

20. Beurel E, Toups M, Nemeroff CB. The Bidirectional Relationship of Depression and Inflammation: Double Trouble. Neuron. 2020;107:234–56.

21. Dantzer R, O’Connor JC, Freund GG, Johnson RW, Kelley KW. From inflammation to sickness and depression: when the immune system subjugates the brain. Nat Rev Neurosci. 2008;9:46–56.

22. Estruch R, Ros E, Salas-Salvadó J, Covas M-I, Corella D, Arós F, et al. Primary Prevention of Cardiovascular Disease with a Mediterranean Diet Supplemented with Extra-Virgin Olive Oil or Nuts. N Engl J Med. 2018;378:e34.

23. Schröder H, Fitó M, Estruch R, Martínez-González MA, Corella D, Salas-Salvadó J, et al. A Short Screener Is Valid for Assessing Mediterranean Diet Adherence among Older Spanish Men and Women. J Nutr. 2011;141:1140–5.

24. Elosua R, Marrugat J, Molina L, Pons S, Pujol E. Validation of the Minnesota Leisure Time Physical Activity Questionnaire in Spanish Men. Am J Epidemiol. 1994;139:1197–209.

25. Elosua R, Garcia M, Aguilar A, Molina L, Covas M-I, Marrugat J. Validation of the Minnesota Leisure Time Physical Activity Questionnaire in Spanish Women: Med Sci Sports Exerc. 2000;32:1431–7.

26. Ribó-Coll M, Castro-Barquero S, Lassale C, Sacanella E, Ros E, Toledo E, et al. Mediterranean Diet and Physical Activity Decrease the Initiation of Cardiovascular Drug Use in High Cardiovascular Risk Individuals: A Cohort Study. Antioxidants. 2021;10:397.

27. Martinez-Gonzalez MA, Corella D, Salas-Salvado J, Ros E, Covas MI, Fiol M, et al. Cohort Profile: Design and methods of the PREDIMED study. Int J Epidemiol. 2012;41:377–85.

28. Castro-Barquero S, Ribó-Coll M, Lassale C, Tresserra-Rimbau A, Castañer O, Pintó X, et al. Mediterranean Diet Decreases the Initiation of Use of Vitamin K Epoxide Reductase Inhibitors and Their Associated Cardiovascular Risk: A Randomized Controlled Trial. Nutrients. 2020;12:3895.

29. Ribó-Coll M, Lassale C, Sacanella E, Ros E, Toledo E, Sorlí JV, et al. Mediterranean diet and antihypertensive drug use: a randomized controlled trial. J Hypertens. 2021;39:1230–7.

30. Schmidt CO, Ittermann T, Schulz A, Grabe HJ, Baumeister SE. Linear, nonlinear or categorical: how to treat complex associations? Splines and nonparametric approaches. Int J Public Health. 2013;58:161–5.

31. Martínez-González MA, Montero P, Ruiz-Canela M, Toledo E, Estruch R, Gómez-Gracia E, et al. Yearly attained adherence to Mediterranean diet and incidence of diabetes in a large randomized trial. Cardiovasc Diabetol. 2023;22:262.

32. Gómez-Pinilla F. Brain foods: the effects of nutrients on brain function. Nat Rev Neurosci. 2008;9:568–78.

33. Bloch MH, Hannestad J. Omega-3 fatty acids for the treatment of depression: systematic review and meta-analysis. Mol Psychiatry. 2012;17:1272–82.

34. Tryfonos C, Pavlidou E, Vorvolakos T, Alexatou O, Vadikolias K, Mentzelou M, et al. Association of Higher Mediterranean Diet Adherence With Lower Prevalence of Disability and Symptom Severity, Depression, Anxiety, Stress, Sleep Quality, Cognitive Impairment, and Physical Inactivity in Older Adults With Multiple Sclerosis. J Geriatr Psychiatry Neurol. 2024;37:318–31.

35. Allcock L, Mantzioris E, Villani A. Adherence to a Mediterranean Diet Is Inversely Associated with Anxiety and Stress but Not Depression: A Cross-Sectional Analysis of Community-Dwelling Older Australians. Nutrients. 2024;16:366.

36. Costa R, Teasdale S, Abreu S, Bastos T, Probst M, Rosenbaum S, et al. Dietary Intake, Adherence to Mediterranean Diet and Lifestyle-Related Factors in People with Schizophrenia. Issues Ment Health Nurs. 2019;40:851–60.

37. Noetel M, Sanders T, Gallardo-Gómez D, Taylor P, Del Pozo Cruz B, van den Hoek D, et al. Effect of exercise for depression: systematic review and network meta-analysis of randomised controlled trials. BMJ. 2024;384:e075847.

38. Gordon BR, McDowell CP, Lyons M, Herring MP. The Effects of Resistance Exercise Training on Anxiety: A Meta-Analysis and Meta-Regression Analysis of Randomized Controlled Trials. Sports Med Auckl NZ. 2017;47:2521–32.

39. Duñabeitia I, Bidaurrazaga-Letona I, Diz JC, Colon-Leira S, García-Fresneda A, Ayán C. Effects of physical exercise in people with epilepsy: A systematic review and meta-analysis. Epilepsy Behav EB. 2022;137:108959.

40. Rißmayer M, Kambeitz J, Javelle F, Lichtenstein TK. Systematic Review and Meta-analysis of Exercise Interventions for Psychotic Disorders: The Impact of Exercise Intensity, Mindfulness Components, and Other Moderators on Symptoms, Functioning, and Cardiometabolic Health. Schizophr Bull. 2024;50:615–30.

41. Hershey MS, Martínez-González MÁ, Álvarez-Álvarez I, Martínez Hernández JA, Ruiz-Canela M. The Mediterranean diet and physical activity: better together than apart for the prevention of premature mortality. Br J Nutr. 2022;128:1413–24.

42. Vuori I. Health benefits of physical activity with special reference to interaction with diet. Public Health Nutr. 2001;4:517–28.

43. Fox KR. The influence of physical activity on mental well-being. Public Health Nutr. 1999;2:411–8.

44. Jacka FN, Berk M. Depression, diet and exercise. Med J Aust [Internet]. 2013 [cited 2024 Oct 4];199. Available from: https://onlinelibrary.wiley.com/doi/abs/10.5694/mja12.10508

45. Fernández-Abascal B, Suárez-Pinilla P, Cobo-Corrales C, Crespo-Facorro B, Suárez-Pinilla M. In- and outpatient lifestyle interventions on diet and exercise and their effect on physical and psychological health: a systematic review and meta-analysis of randomised controlled trials in patients with schizophrenia spectrum disorders and first episode of psychosis. Neurosci Biobehav Rev. 2021;125:535–68.

